# Timing of Addiction Consultation and Alcohol Withdrawal Management in Hospitalized Adults

**DOI:** 10.64898/2026.10.18.26365824

**Authors:** Thomas J. Reese, Kristopher A. Kast, David Marcovitz, Lisa Stewart-Kast, Hilary A. Tindle, Adam Wright, Mauli V. Shah, Sahit N. Menon, Amanda von Horn

## Abstract

**Objective:** To evaluate the association of early addiction consultation with withdrawal pharmacotherapy and complications in hospitalized patients with alcohol withdrawal.

**Methods:** Retrospective single-center cohort of adult hospitalizations with alcohol withdrawal (diagnosis code or CIWA-Ar ≥10 within 24 hours) from July 2020 to January 2026. Exposure was an addiction consult service order within 24 hours of arrival, compared with no or later consultation. Primary outcomes were intensive care unit (ICU) transfer after the first 24 hours and delirium tremens or withdrawal seizure not present on admission. Process outcomes included phenobarbital and thiamine administration within 24 hours and benzodiazepine dose in the first 48 hours. Secondary outcomes included length of stay and 30-day death or return to hospital.

**Results:** Of 4,663 hospitalizations, 1,379 (29.6%) received early consultation. After adjustment, early consultation was associated with lower odds of ICU transfer (odds ratio [OR] 0.73; 95% CI 0.54–0.99) and fewer episodes of delirium tremens (risk difference −1.09 per 100; −1.45 to −0.73), with no significant difference in withdrawal seizure. Early consultation was associated with phenobarbital (OR 1.57; 1.32–1.86) and thiamine (OR 1.23; 1.07–1.41) within 24 hours, higher 48-hour lorazepam-equivalent dose (ratio of geometric means 1.44; 1.32–1.57), shorter stays (ratio 0.92; 0.88–0.97), and lower odds of 30-day death or return to hospital (OR 0.78; 0.66–0.91).

**Conclusions:** Early addiction consultation was associated with earlier, more intensive withdrawal pharmacotherapy, lower odds of ICU transfer, and fewer episodes of delirium tremens, although the complication associations were sensitive to cohort definition.

## INTRODUCTION

Alcohol withdrawal complicates more than 500,000 hospitalizations annually in the United States and can cause seizures, delirium tremens, and intensive care unit (ICU) admission.^1–3^ Guidelines recommend symptom-triggered benzodiazepine therapy, early thiamine, and structured monitoring, yet inpatient care is characterized by variable benzodiazepine use, delayed thiamine, and limited uptake of medications for alcohol use disorder (MAUD), which fewer than 5% of eligible patients receive.^1,4–9^ These gaps are associated with ICU transfer, longer stays, and readmission.^7,10^ Early, adequate dosing and adjunctive agents such as phenobarbital may prevent escalation to severe withdrawal, but their use varies across clinicians.^11–15^

Inpatient addiction consultation services (ACS) provide assessment of substance use disorders, withdrawal management, MAUD initiation, counseling, and linkage to treatment.^16,17^ Among adults hospitalized with alcohol use disorder, ACS consultation has been associated with more MAUD prescribing and, in some settings, less subsequent acute care use, with mixed effects on mortality and readmission.^17–20^ Prior evaluations rarely examined withdrawal-specific processes such as benzodiazepine dosing, phenobarbital use, and thiamine timing, and most defined exposure as consultation at any point during the stay. Most involved heterogeneous substance use populations and defined exposure as consultation at any point during the stay.^17–19,21^ Whether ACS changes withdrawal-specific care and short-term outcomes, and whether the timing of consultation matters, remains uncertain.

In a prior study, ACS consultation was associated with lower odds of death or return to hospital within 30 days, but only about 20% of this association was mediated by discharge MAUD.^22^ We hypothesized that management of acute withdrawal during the first day is a principal pathway, and examined whether ACS consultation within 24 hours of arrival was associated with withdrawal pharmacotherapy and in-hospital complications, comparing early with later consultation to separate timing from consultation itself.

## METHODS

### Study Design and Setting

We conducted a retrospective cohort study at Vanderbilt University Hospital, using electronic health record data from January 2018 to January 2026. The institutional review board at Vanderbilt University Medical Center approved the study with a waiver of informed consent. We followed the Strengthening the Reporting of Observational Studies in Epidemiology (STROBE) reporting guideline (**Supplementary Table 3**).^23^

The ACS is a multidisciplinary team of addiction physicians, advanced practice clinicians, and social workers that provides daytime consultation seven days a week to all inpatient services. Although the consult order existed earlier, the service was formally staffed and promoted to admitting teams in January 2021. Throughout the study period, withdrawal management on all services followed an institutional order set aligned with national guidelines: CIWA-Ar monitoring with symptom-triggered benzodiazepine dosing, scheduled thiamine, and phenobarbital available at clinician discretion as an adjunct or alternative for severe or benzodiazepine-refractory withdrawal. The order set did not change substantively during the study period, and consultants used the same agents.

### Participants

We identified adult (≥18 years) inpatient hospitalizations with evidence of alcohol withdrawal, defined as either an alcohol withdrawal diagnosis (ICD-10, F10.13x or F10.23x) or a maximum Clinical Institute Withdrawal Assessment for Alcohol, revised (CIWA-Ar) score of 10 or higher within 24 hours of arrival. We excluded hospitalizations shorter than 24 hours, deaths within 24 hours, and admissions to the psychiatric hospital. Because consultation for alcohol withdrawal increased before formalization, we defined a steady-state period empirically as beginning with the first of three consecutive quarters in which the proportion of withdrawal hospitalizations receiving early consultation was at least 80% of the median over the final eight study quarters. The primary cohort comprised confirmed withdrawal hospitalizations in the steady-state period (July 2020 to January 2026). Hospitalizations with lower CIWA-Ar scores or before July 2020 were included in sensitivity analyses.

### Exposure

We defined time zero as emergency department (ED) arrival for patients admitted through the ED and inpatient admission otherwise, so that ED medications and CIWA-Ar scores were attributed to the hospitalization. Exposure was an ACS consult order placed within 24 hours of time zero (early consultation). We used the order time because documentation of consult completion lags the clinical encounter. The comparison group comprised hospitalizations with no or later consultation; the latter were examined separately in a timing analysis.

### Outcomes

Our primary outcomes were withdrawal-specific in-hospital complications: (1) ICU transfer from a non-ICU service after a 24-hour landmark, (2) delirium tremens (F10.231 or F10.131) not present on admission, and (3) withdrawal seizure (R56.9 or G40.5x) not present on admission.^24^ Diagnoses with a present-on-admission indicator of yes, or recorded only on the ED or encounter diagnosis list, were classified as present on admission.^25^ Diagnoses with an undetermined or exempt indicator (12 delirium tremens and 25 seizure) counted as neither. Delirium tremens and seizure present on admission were treated as baseline covariates because they precede and commonly prompt consultation.

Process outcomes were withdrawal pharmacotherapy, identified from medication administration records after time zero: any benzodiazepine; lorazepam-equivalent dose among benzodiazepine recipients in the first 48 hours, first 72 hours, per hospital day, and for the whole stay; phenobarbital and thiamine, at any time and within 24 hours; and adjunctive agents (clonidine, dexmedetomidine, gabapentin). We reported midazolam separately.

Secondary outcomes were length of stay; discharge disposition (routine, patient-directed discharge [PDD], in-hospital death); all-cause readmission, ED visit, and death within 30 days of discharge, and their composite (death or return to hospital), among patients discharged alive; and inpatient MAUD initiation and new MAUD at discharge (naltrexone, acamprosate, or disulfiram) among patients without MAUD on the admission medication list.

### Covariates

Covariates, measured at or before hospitalization except CIWA-Ar variables, were age, sex, race, Hispanic ethnicity, and insurance; initial CIWA-Ar score, maximum score within 24 hours, and absence of any score during the stay (missing scores set to zero with an indicator); delirium tremens and seizure present on admission; admission from the ED to an ICU; delirium tremens, withdrawal seizure, and withdrawal hospitalization in the preceding three years; and opioid use disorder, other substance use disorders, liver disease, heart failure, seizure disorder, depressive disorder, psychotic disorder, and the Elixhauser comorbidity count.^26^ Prior use comprised MAUD on the admission medication list and admissions and ED visits in the prior 12 months; hospitalization characteristics were admission year and weekend admission. Covariates were complete except insurance; hospitalizations without a coverage record (8%) were classified as self-pay/other.

### Statistical Analysis

We compared baseline characteristics using standardized mean differences (SMDs; >0.10 indicating imbalance), summarized variables as counts (percentages) or medians (interquartile ranges), and compared unadjusted outcomes with χ² tests.

We estimated a propensity score for early consultation by logistic regression on all covariates and applied overlap weighting, which weights each hospitalization by the probability of the other treatment, targets the population with substantial probability of either early consultation or usual care, and yields exact mean balance on the propensity covariates.^27,28^ Because patients could contribute more than one hospitalization, standard errors were clustered on patient. We re-estimated weights in each analytic subset except for outcomes among patients discharged alive, which used the full-cohort weights

For primary outcomes, we analyzed ICU transfer among hospitalizations not in an ICU during the first 24 hours, with follow-up from the 24 hour landmark, to exclude transfers within the exposure window that preclude consultation rather than result from it.^24^ We fit overlap-weighted logistic regression adjusting for withdrawal severity, admission from the ED to ICU, complications present on admission, prior withdrawal history, Elixhauser count, liver disease, opioid use disorder, and prior utilization, and report odds ratios (ORs) with 95% CIs. We also fit a cause-specific Cox model from the landmark with discharge as a competing event. For delirium tremens and seizure not present on admission, which had fewer than 10 exposed events, we report the overlap-weighted risk difference (RD) with patient-clustered delta-method confidence intervals (CIs), without an outcome model.

Binary process outcomes used the same overlap-weighted logistic models. Benzodiazepine dose was analyzed as a two-part model: any administration, then log-transformed lorazepam-equivalent dose among recipients, reported as ratios of geometric means (RGMs). Because 24-hour process outcomes could precede the consult order, we described time to the order and the proportion of first phenobarbital and thiamine doses given before it.

We analyzed log-transformed length of stay with overlap-weighted linear models (RGM); discharge disposition with cause-specific Cox models from admission treating other dispositions as competing events (hazard ratios, HRs); and 30-day and MAUD outcomes with overlap-weighted logistic models among patients discharged alive.

We performed three sets of additional analyses. First, to separate timing from consultation, we compared early with later consultation among consulted hospitalizations, re-estimating weights; because a later consult requires remaining in hospital beyond 24 hours, inference was restricted to process outcomes in fixed early windows. Second, we repeated the core analyses in six alternative cohorts; with exposure redefined as consultation within 48 hours; in a 1:1 propensity-matched cohort (caliper 0.2 SD of the logit); with adjustment for palliative care consultation within 24 hours as an acuity marker; among hospitalizations without delirium tremens or seizure at admission; with entry restricted to a maximum CIWA-Ar of 10 or higher within 24 hours, which removes entry by diagnosis code; and omitting the 24-hour maximum CIWA-Ar, which is measured within the exposure window. We calculated E-values for the primary outcomes.^29^ Third, we re-estimated weights within subgroups defined by withdrawal severity (maximum CIWA-Ar within 24 hours <10, 10–18, ≥19), prior utilization (≥3 ED visits or ≥2 admissions in 12 months), and opioid use disorder, without tests of interaction, and described outcomes stratified by PDD. Analyses used R 4.3, 2-sided α = .05, and 95% CIs without adjustment for multiple comparisons.

## RESULTS

Among 12,913 hospitalizations with evidence of alcohol withdrawal over the full study period, 8,796 (68%) were identified by CIWA-Ar monitoring alone, with a median maximum score of 6 and early consultation in 8%. Early consultation rose from 0% in early 2018 to about 20% by the third quarter of 2020 and remained stable through 2025 (**Figure 1**). The primary cohort comprised 4,663 hospitalizations of 3,365 patients, of which 1,379 (29.6%) received ACS consultation within 24 hours and 3,284 did not, including 832 consulted later in the stay (**Figure 2**). Patients with early ACS were younger and more often had severe withdrawal, complications present on admission, prior withdrawal hospitalization, and opioid use disorder; after overlap weighting, all covariates were balanced (effective sample size 2,354 unexposed, 1,267 exposed; **Table 1**).

**Figure 1.**
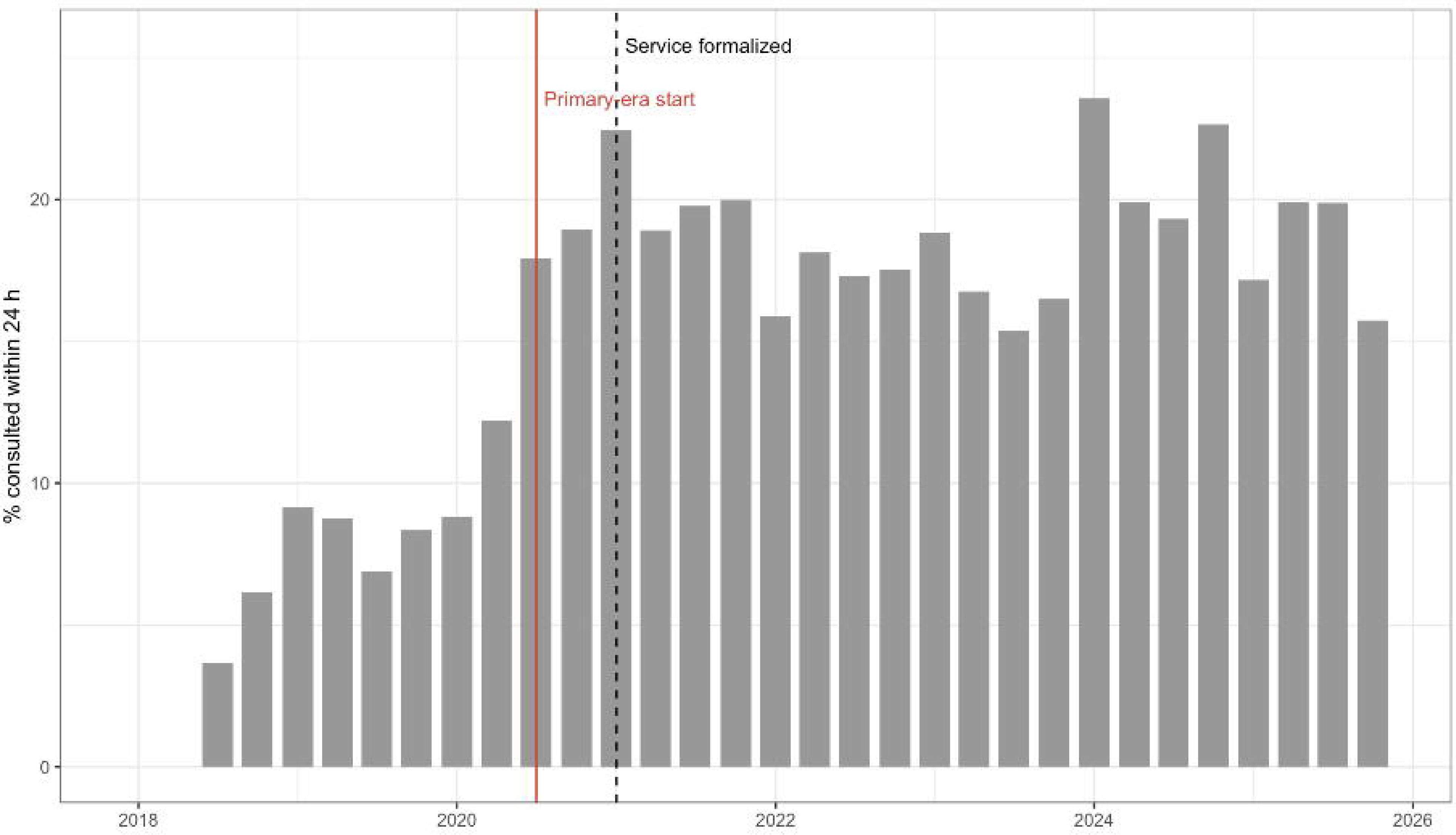
Proportion of alcohol withdrawal hospitalizations receiving an addiction consult order within 24 hours of arrival, by calendar quarter, 2018–2025. The dashed line marks formal staffing of the service (January 2021); the solid line marks the empirically defined start of the steady-state period (July 2020) used for the primary analysis.

**Figure 2.**
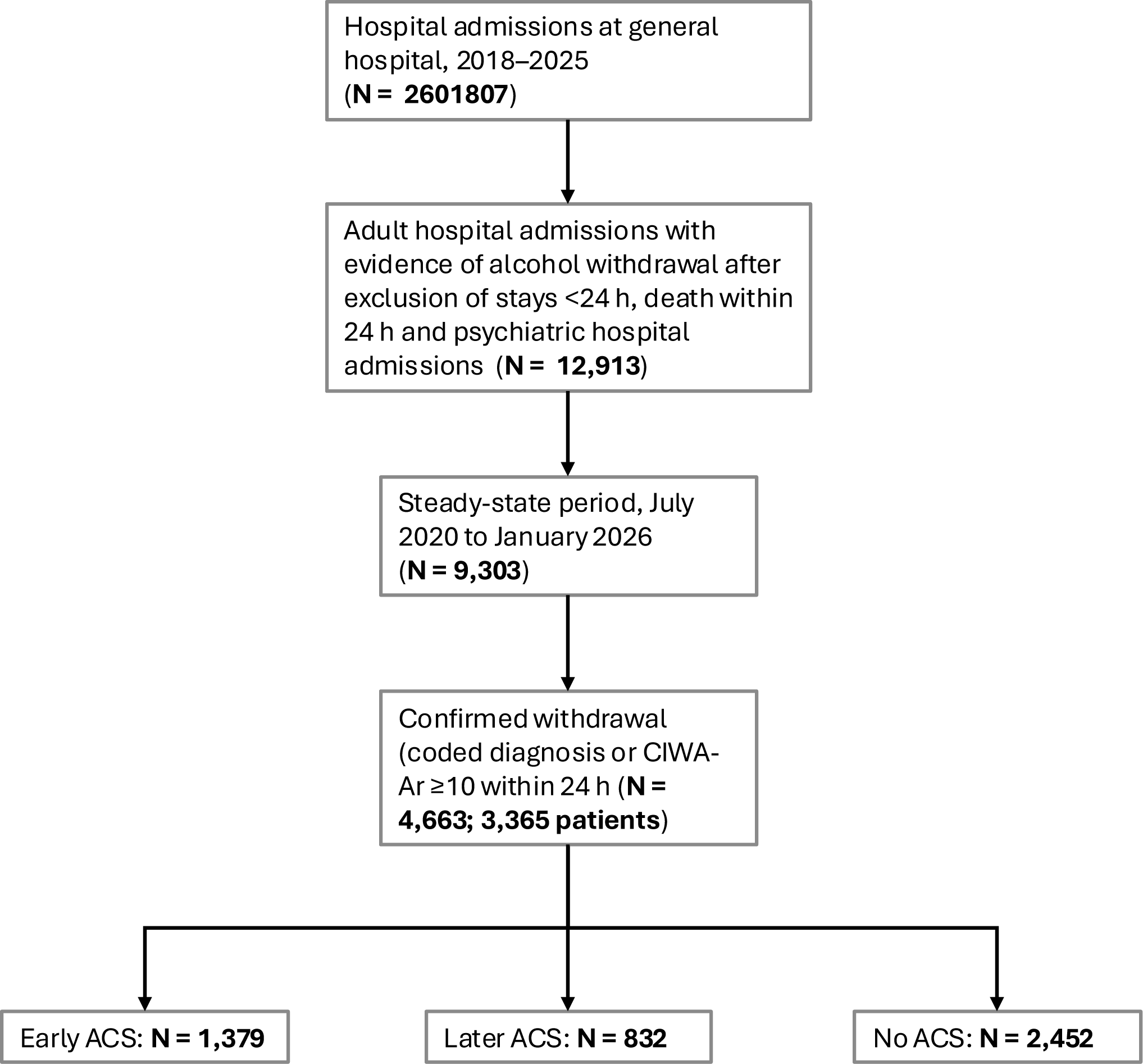
Study flow diagram.

**Table 1.** Baseline characteristics of hospitalizations with confirmed alcohol withdrawal, by early ACS consultation (primary cohort, n = 4663)

| Characteristic | No early ACS<br>(N = 3284) | Early ACS<br>(N = 1379) | SMD |
| --- | --- | --- | --- |
| **Demographics** |  |  |  |
| Age, y, median (IQR) | 50 (39-60) | 43 (35-54) | 0.38 |
| Male | 2486 (75.7) | 1010 (73.2) | 0.06 |
| **Race** |  |  | 0.07 |
| White | 2665 (81.2) | 1153 (83.6) |  |
| Black | 457 (13.9) | 172 (12.5) |  |
| Other or unknown | 162 (4.9) | 54 (3.9) |  |
| **Hispanic** | 125 (3.8) | 53 (3.8) | 0.00 |
| **Insurance** |  |  | 0.21 |
| Commercial | 1236 (37.6) | 584 (42.3) |  |
| Medicare or government | 972 (29.6) | 286 (20.7) |  |
| Medicaid | 772 (23.5) | 396 (28.7) |  |
| Self-pay or other | 304 (9.3) | 113 (8.2) |  |
| **Withdrawal Severity and Presentation** |  |  |  |
| Initial CIWA-Ar score, median (IQR) | 11 (7–18) | 13 (7–21) | 0.18 |
| Maximum CIWA-Ar within 24 h, median (IQR) | 17 (11–23) | 20 (13–26) | 0.26 |
| No CIWA-Ar score within 24 h | 528 (16.1) | 50 (3.6) | 0.31 |
| Delirium tremens present on admission | 214 (6.5) | 138 (10.0) | 0.13 |
| Seizure present on admission | 304 (9.3) | 181 (13.1) | 0.12 |
| ICU admission from ED | 525 (16.0) | 97 (7.0) | 0.28 |
| **History (prior 3 years)** |  |  |  |
| Delirium tremens | 267 (8.1) | 185 (13.4) | 0.17 |
| Withdrawal seizure | 367 (11.2) | 209 (15.2) | 0.12 |
| Withdrawal hospitalization | 997 (30.4) | 651 (47.2) | 0.35 |
| **Comorbid Conditions** |  |  |  |
| Opioid use disorder | 443 (13.5) | 466 (33.8) | 0.49 |
| Other substance use disorder | 2056 (62.6) | 1040 (75.4) | 0.28 |
| Liver disease | 696 (21.2) | 372 (27.0) | 0.14 |
| Heart failure | 395 (12.0) | 149 (10.8) | 0.04 |
| Depressive disorder | 601 (18.3) | 418 (30.3) | 0.28 |
| Psychotic disorder | 559 (17.0) | 328 (23.8) | 0.17 |
| Elixhauser count, median (IQR) | 5 (3–7) | 5 (4–7) | 0.13 |
| **Prior Health Care Use (12 months)** |  |  |  |
| Hospital admissions, median (IQR) | 0 (0–1) | 0 (0–2) | 0.22 |
| ED visits, median (IQR) | 0 (0–2) | 1 (0–4) | 0.22 |
| Home medication for AUD at admission | 85 (2.6) | 67 (4.9) | 0.12 |
| **Hospitalization** |  |  |  |
| Surgical admitting service | 530 (16.1) | 48 (3.5) | 0.44 |
| Weekend admission | 1087 (33.1) | 363 (26.3) | 0.15 |

Values are n (%) unless stated. Admission year was included in the propensity model (not shown). After overlap weighting, all SMDs were 0.00. Abbreviations: ACS, Addiction Consultation Service; CIWA-Ar, Clinical Institute Withdrawal Assessment for Alcohol, revised; ED, emergency department; ICU, intensive care unit; IQR, interquartile range; MAUD, medication for alcohol use disorder; SMD, standardized mean difference.

### Unadjusted Outcomes

Early ACS hospitalizations less often had ICU transfer after 24 hours (6.5% vs 8.5%), delirium tremens not present on admission (2 [0.1%] vs 64 [1.9%]), and withdrawal seizure not present on admission (4 [0.3%] vs 27 [0.8%]), but more often had these complications present on admission; 352 of 430 coded delirium tremens hospitalizations (82%) were present on admission (**Table 2**). They received higher 48-hour lorazepam-equivalent doses (median 10 vs 4 mg), more phenobarbital and thiamine within 24 hours, and less dexmedetomidine and midazolam, and had shorter stays, lower mortality, and more PDD and MAUD initiation.

**Table 2.** Unadjusted withdrawal management, complications and outcomes by early ACS consultation (primary cohort)

| **Measure** | **No early ACS**<br>**(N = 3284)** | **Early ACS**<br>**(N = 1379)** | **P Value** |
| --- | --- | --- | --- |
| **Withdrawal complications** |  |  |  |
| ICU transfer after 24 hours<sup>a</sup> | 196 (8.5) | 75 (6.5) | .04 |
| Delirium tremens, present on admission | 214 (6.5) | 138 (10.0) | <.001 |
| Delirium tremens, not present on admission | 64 (1.9) | 2 (0.1) | <.001 |
| Withdrawal seizure, present on admission | 304 (9.3) | 181 (13.1) | <.001 |
| Withdrawal seizure, not present on admission | 27 (0.8) | 4 (0.3) | .07 |
| **Withdrawal pharmacotherapy** |  |  |  |
| Any benzodiazepine | 2021 (61.5) | 1054 (76.4) | <.001 |
| Lorazepam-eq dose, first 48 hours, mg, median (IQR)<sup>b</sup> | 4 (2–12) | 10 (4–18) | — |
| Lorazepam-eq dose, whole stay, mg, median (IQR)<sup>b</sup> | 6 (2–14) | 12 (4–22) | — |
| Phenobarbital within 24 hours | 665 (20.2) | 385 (27.9) | <.001 |
| Phenobarbital, any | 962 (29.3) | 450 (32.6) | .03 |
| Benzodiazepine and phenobarbital | 525 (16.0) | 339 (24.6) | <.001 |
| Thiamine within 24 hours | 1462 (44.5) | 723 (52.4) | <.001 |
| Thiamine, any | 2203 (67.1) | 982 (71.2) | .006 |
| Clonidine | 253 (7.7) | 198 (14.4) | <.001 |
| Dexmedetomidine | 427 (13.0) | 59 (4.3) | <.001 |
| Midazolam | 497 (15.1) | 108 (7.8) | <.001 |
| Gabapentin | 986 (30.0) | 429 (31.1) | .48 |
| **Hospitalization and 30-day outcomes** |  |  |  |
| Length of stay, d, median (IQR) | 4.5 (2.5–8.9) | 3.9 (2.6–6.2) | — |
| Patient-directed discharge | 188 (5.7) | 136 (9.9) | <.001 |
| In-hospital death | 114 (3.5) | 11 (0.8) | <.001 |
| 30-day readmission<sup>c</sup> | 593 (18.7) | 285 (20.8) | .11 |
| 30-day ED visit<sup>c</sup> | 819 (25.8) | 371 (27.1) | .39 |
| 30-day death<sup>c</sup> | 55 (1.7) | 2 (0.1) | <.001 |
| 30-day death or return to hospital<sup>c</sup> | 965 (30.4) | 410 (30.0) | .78 |
| Inpatient MAUD initiation | 396 (12.1) | 417 (30.2) | <.001 |
| New MAUD at discharge<sup>c</sup> | 327 (10.0) | 340 (24.7) | <.001 |

Values are n (%) unless stated. *P* values from $\chi^2$ tests, or Fisher's exact test for outcomes with fewer than 10 events in either group; medians are descriptive. <sup>a</sup>Among 3,431 hospitalizations not in an ICU during the first 24 hours. <sup>b</sup>Among hospitalizations receiving any benzodiazepine. <sup>c</sup>Among 4,538 hospitalizations discharged alive. Abbreviations: ACS, Addiction Consultation Service; ED, emergency department; eq, equivalent; ICU, intensive care unit; IQR, interquartile range; MAUD, medication for alcohol use disorder.

### Adjusted Outcomes

In weighted models, early ACS consultation was associated with lower odds of ICU transfer after 24 hours (OR 0.73; 95% CI 0.54–0.99; cause-specific HR 0.76; 0.57–1.01), fewer episodes of delirium tremens not present on admission (RD −1.09 per 100 hospitalizations; −1.45 to −0.73; RR 0.08; 0.02–0.32), and a nonsignificant reduction in withdrawal seizure not present on admission (RD −0.34 per 100; −0.73 to 0.05) (**Table 3**). Defining ICU transfer as any entry after 6 hours strengthened the association (OR 0.56; 0.46–0.69), but 568 of 839 such transfers (68%) occurred within the exposure window and 251 preceded the consult order; any coded delirium tremens, including present on admission, showed no association (OR 0.91; 0.72–1.16).

**Table 3.**
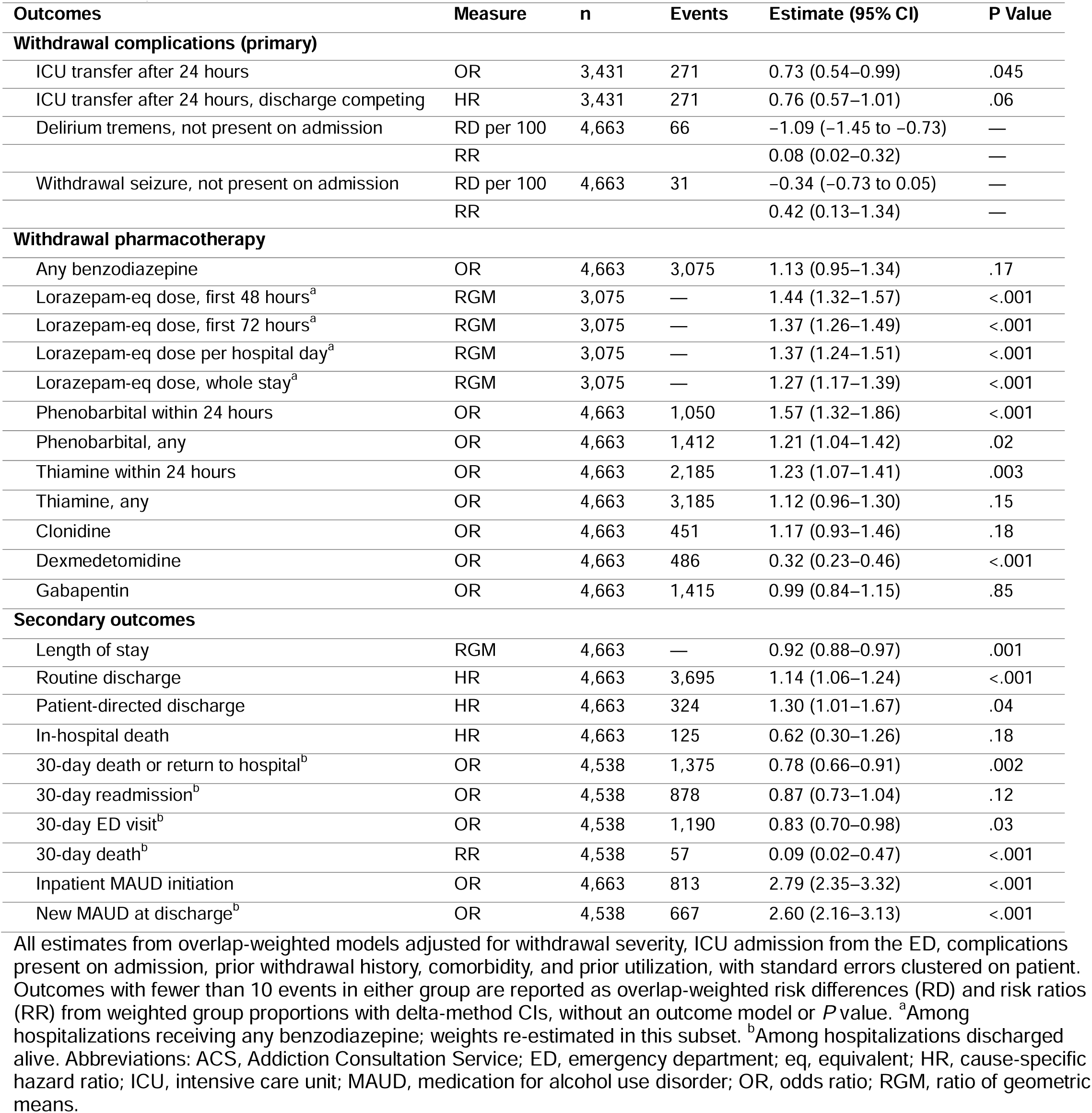
Adjusted associations of early ACS consultation with withdrawal management, complications and outcomes (primary cohort).

Early ACS was associated with higher lorazepam-equivalent dose among benzodiazepine recipients in the first 48 hours (RGM 1.44; 1.32–1.57) but not with any benzodiazepine use (OR 1.13; 0.95–1.34); with phenobarbital within 24 hours (OR 1.57; 1.32–1.86) and thiamine within 24 hours (OR 1.23; 1.07–1.41); and with lower odds of dexmedetomidine (OR 0.32; 0.23–0.46). Clonidine and gabapentin did not differ. Early ACS was also associated with inpatient MAUD initiation (OR 2.79; 2.35–3.32) and new MAUD at discharge (OR 2.60; 2.16–3.13).

Early ACS was associated with shorter length of stay (RGM 0.92; 0.88–0.97), higher rates of routine discharge (HR 1.14; 1.06–1.24) and PDD (HR 1.30; 1.01–1.67), and no difference in in-hospital mortality. Among patients discharged alive, early ACS was associated with lower odds of the 30-day composite (OR 0.78; 0.66–0.91) and ED visit (OR 0.83; 0.70–0.98) but not readmission; 30-day death occurred in 2 vs 55 patients.

### Additional Analyses

Among 2,211 hospitalizations consulted at any time, early compared with later consultation was associated with phenobarbital within 24 hours (OR 1.61; 1.26–2.06), higher 48-hour lorazepam-equivalent dose (RGM 1.29; 1.15–1.46), and thiamine within 24 hours (OR 1.25; 1.04–1.51), but not with any phenobarbital or any benzodiazepine use. Among early consultations, the order was placed a median of 14.1 hours (IQR 9.5–18.5) after time zero, and 60.5% of first phenobarbital and 69.3% of first thiamine doses within 24 hours preceded it. Process, length-of-stay, 30-day, and delirium tremens associations were consistent across the six alternative cohorts, and the ICU transfer OR ranged from 0.61 to 0.85 (**Supplementary Table 1**). Estimates were similar with a 48-hour exposure window, in the matched cohort, with adjustment for palliative care consultation, among hospitalizations without delirium tremens at admission (RD −1.20 per 100; −1.60 to −0.81), and without the 24-hour maximum CIWA-Ar (ICU transfer OR 0.71; 0.52–0.96); E-values were 2.1 for ICU transfer and 1.5 for the 30-day composite (**Supplementary Table 2**). Restricting entry to a maximum CIWA-Ar of 10 or higher within 24 hours, which excluded 330 hospitalizations entered by diagnosis code alone (9.2% of unexposed vs 2.1% of exposed), left process, length-of-stay, and 30-day associations unchanged but attenuated ICU transfer (OR 0.82; 0.58–1.15) and delirium tremens (RR 0.24; 0.05–1.08). Phenobarbital associations were larger in severe withdrawal (OR 1.48; 1.20–1.83) and among higher utilizers (OR 1.51; 1.16–1.97), as was the 30-day composite in severe withdrawal (OR 0.74; 0.59–0.93). PDD hospitalizations had higher 30-day event rates regardless of consultation (40–45% vs 28–30%), but those with early ACS more often received phenobarbital (39.7% vs 24.5%) and early thiamine (53.7% vs 44.1%).

## DISCUSSION

Among adults hospitalized with alcohol withdrawal, ACS consultation within 24 hours of arrival was associated with earlier and more intensive withdrawal treatment: more phenobarbital, higher 48-hour benzodiazepine dosing, earlier thiamine, and less dexmedetomidine. Early consultation was also associated with lower odds of ICU transfer after the first day and fewer episodes of delirium tremens arising after admission. Process, length-of-stay, and 30-day findings were robust across sensitivity analyses, whereas the ICU transfer and delirium tremens associations were attenuated and no longer significant when cohort entry required CIWA-Ar confirmation.

These results address the question left open by our previous study, in which discharge MAUD explained little of the association between consultation and 30 day outcomes.^22^ Withdrawal management is a plausible pathway, although the evidence here is indirect. Consultants at our center favor front-loaded benzodiazepine dosing and early phenobarbital for severe or escalating withdrawal.^11–15^ Most first phenobarbital and thiamine doses among early consultations, however, preceded the consult order, so ED and admitting teams contributed to the 24-hour differences, and early consultation may partly mark patients already receiving guideline-concordant care. Two findings nonetheless point to consultation itself: higher benzodiazepine dosing over 48 hours, which extends past most consult orders, and the early-versus-later comparison, in which earlier consultation was associated with earlier phenobarbital and thiamine and higher early dosing but not with whether these drugs were used.

Outcome definitions matter for evaluations of consult services. Two thirds of ICU transfers measured from admission occurred within the first 24 hours and many preceded the consult order. Early deterioration reduces the chance of consultation, and counting those events overstates benefit.^24^ Additionally, most coded delirium tremens was present on admission and more common among consulted patients, so any coded delirium tremens showed no association whereas delirium tremens not present on admission did.

Selection also shapes the mortality and PDD results. Patients who are moribund, intubated, or receiving comfort care on the first day are less likely to be consulted and more likely to die, so the very low mortality among consulted patients likely reflects selection rather than consultation. Consultation was associated with a higher rate of PDD, as in prior studies.^30,31^ Consulted patients who left by PDD had received more phenobarbital and thiamine than non-consulted PDD patients and had no worse 30-day outcomes, consistent with consultation reaching patients at high baseline risk of leaving rather than causing departures.^31,32^

Prior evaluations examined heterogeneous substance use populations, defined exposure as any consultation, and reported MAUD prescribing and post-discharge utilization rather than withdrawal specific processes and complications.^18–21,30–37^ Here, early consultation also more than doubled the odds of MAUD initiation, as in our prior study, and reinforced guideline-recommended withdrawal management in general medical settings; the larger phenobarbital and 30-day estimates in severe withdrawal and among high utilizers suggest prioritizing these patients when consult capacity is limited.^15,38,39^

### Limitations

This single-center study reflects one service, its referral patterns, and local withdrawal protocols. Cohort entry relied on CIWA-Ar monitoring, which is not applied uniformly across services (16% of unexposed hospitalizations had no score within 24 hours), and on diagnosis codes assigned at discharge; hospitalizations entered by code alone were predominantly unexposed and may include patients too ill to be scored or consulted, and without them the ICU transfer and delirium tremens associations were no longer statistically significant. As noted, the 24-hour process associations partly reflect admitting-team treatment that preceded the consult order. Residual confounding by clinician selection remains possible: E-values indicate that an unmeasured confounder associated with exposure and outcome by a risk ratio of about 2 could explain the ICU transfer association, and the only acuity marker we could capture, palliative care consultation, was rare; ventilation and code-status orders were not retrievable. Delirium tremens and seizure were ascertained from diagnosis codes, and present-on-admission assignment depends on documentation: a consultant may document emerging delirium on the first day that is coded as present on admission, whereas the same delirium in a non-consulted patient would be coded as hospital-acquired, inflating the association; this cannot be excluded without chart review. Subgroup differences were not formally tested. Readmissions and deaths outside the health system were not captured. Process measures describe the intensity and timing of therapy, not its appropriateness, and we did not assess oversedation or other adverse effects.

## CONCLUSION

Among hospitalized adults with alcohol withdrawal, addiction consultation within 24 hours of arrival was associated with earlier, more intensive withdrawal pharmacotherapy, shorter hospitalizations, lower odds of ICU transfer, and fewer episodes of delirium tremens, although the complication associations were sensitive to cohort definition. Withdrawal management during the first day is a plausible, though not fully isolated, pathway for the short-term benefits of consultation that discharge pharmacotherapy does not explain.

## Data Availability

The data are not available due to privacy or ethical restrictions.

## ACKNOWLEDGEMENTS

Contributors: The authors thank the Vanderbilt University Medical Center Addiction Consult Service clinicians and staff for their dedication to patient care and their contributions to the development of the service evaluated in this study.

## Author Contribution

Thomas J. Reese contributed to study conception and design, data acquisition, data analysis and interpretation, and drafting and critical revision of the manuscript. Kristopher A. Kast contributed to study conception and design, interpretation of findings, and drafting and critical revision of the manuscript. David Marcovitz contributed to study conception and design, interpretation of findings, and critical revision of the manuscript. Lisa Stewart-Kast contributed to interpretation of findings and critical revision of the manuscript. Hilary A. Tindle contributed to interpretation of findings and critical revision of the manuscript. Adam Wright contributed to interpretation of findings and critical revision of the manuscript. Mauli V. Shah contributed to data acquisition and critical revision of the manuscript. Sahit N. Menon contributed to interpretation of findings and critical revision of the manuscript. Amanda von Horn contributed to study conception and design, interpretation of findings, and critical revision of the manuscript. All authors approved the final manuscript.

