## Supplement Tables 1, 2, 3 for "Timing of Addiction Consultation and Alcohol Withdrawal Management in Hospitalized Adults"

| **Supplementary Table 1. Core estimates across cohort definitions (propensity scores and overlap weights re-estimated within each cohort).** | | | | | | | |
| --- | --- | --- | --- | --- | --- | --- | --- |
| Outcome | Primary: steady state + confirmed withdrawal (n = 4,663) | Steady state, all (n = 9,303) | Formal period + confirmed (n = 4,247) | Full period + confirmed (n = 6,328) | Full period, all (n = 12,913) | Coded withdrawal only (n = 3,153) | Non-surgical services (n = 4,085) |
| ICU transfer after 24 hours, OR | 0.73 (0.54–0.99) | 0.67 (0.51–0.89) | 0.76 (0.56–1.04) | 0.65 (0.48–0.87) | 0.61 (0.47–0.79) | 0.63 (0.45–0.88) | 0.85 (0.61–1.17) |
| DT not present on admission, RR | 0.08 (0.02–0.32) | 0.09 (0.02–0.37) | 0.10 (0.02–0.41) | 0.20 (0.07–0.58) | 0.26 (0.09–0.77) | 0.05 (0.01–0.21) | 0.10 (0.02–0.44) |
| Seizure not present on admission, RR | 0.42 (0.13–1.34) | 0.54 (0.19–1.57) | 0.43 (0.13–1.40) | 0.35 (0.11–1.07) | 0.42 (0.15–1.15) | 0.61 (0.19–1.96) | 0.83 (0.25–2.79) |
| Any benzodiazepine, OR | 1.13 (0.95–1.34) | 1.15 (1.00–1.33) | 1.12 (0.94–1.34) | 1.12 (0.96–1.32) | 1.12 (0.98–1.28) | 0.76 (0.61–0.94) | 1.07 (0.89–1.28) |
| Lorazepam-eq dose, recipients, RGM | 1.27 (1.17–1.39) | 1.28 (1.18–1.39) | 1.24 (1.13–1.35) | 1.22 (1.13–1.32) | 1.22 (1.13–1.31) | 1.11 (1.01–1.21) | 1.25 (1.15–1.36) |
| Phenobarbital, any, OR | 1.21 (1.04–1.42) | 1.28 (1.11–1.49) | 1.21 (1.03–1.42) | 1.24 (1.07–1.44) | 1.35 (1.18–1.56) | 0.98 (0.83–1.17) | 1.52 (1.29–1.81) |
| Thiamine within 24 hours, OR | 1.23 (1.07–1.41) | 1.29 (1.15–1.46) | 1.20 (1.04–1.39) | 1.21 (1.06–1.37) | 1.25 (1.12–1.39) | 1.15 (0.98–1.35) | 1.31 (1.14–1.51) |
| Length of stay, RGM | 0.92 (0.88–0.97) | 0.90 (0.86–0.94) | 0.92 (0.87–0.96) | 0.93 (0.89–0.97) | 0.90 (0.87–0.94) | 0.88 (0.84–0.93) | 0.95 (0.91–1.00) |
| 30-day death or return to hospital, OR | 0.78 (0.66–0.91) | 0.80 (0.70–0.92) | 0.81 (0.68–0.96) | 0.76 (0.66–0.89) | 0.79 (0.69–0.90) | 0.82 (0.68–0.98) | 0.78 (0.66–0.92) |
| Inpatient MAUD initiation, OR | 2.79 (2.35–3.32) | 3.22 (2.74–3.78) | 2.73 (2.28–3.27) | 2.80 (2.38–3.30) | 3.28 (2.82–3.81) | 2.16 (1.80–2.61) | 2.68 (2.25–3.19) |

“Steady state” is July 2020 onward; “formal period” is January 2021 onward; “confirmed withdrawal” is a coded withdrawal diagnosis or CIWA-Ar ≥10 within 24 hours; “all” includes hospitalizations identified by CIWA-Ar monitoring alone with a maximum score <10. Abbreviations: DT, delirium tremens; eq, equivalent; MAUD, medication for alcohol use disorder; OR, odds ratio; RGM, ratio of geometric means; RR, overlap-weighted risk ratio.

| **Supplementary Table 2. Additional analyses in the primary cohort.** | | | |
| --- | --- | --- | --- |
| **Analysis** | | **Outcome** | **Estimate (95% CI)** |
| **Early (≤24 hours) vs later consultation, among 2,211 hospitalizations consulted at any time (1,379 early, 832 later)** | | | |
|  | Fixed-window process outcomes | Any benzodiazepine, OR | 0.86 (0.67–1.10) |
|  |  | Lorazepam-eq dose, first 48 hours, recipients, RGM | 1.29 (1.15–1.46) |
|  |  | Phenobarbital within 24 hours, OR | 1.61 (1.26–2.06) |
|  |  | Phenobarbital, any, OR | 1.00 (0.81–1.24) |
|  |  | Thiamine within 24 hours, OR | 1.25 (1.04–1.51) |
|  | Exploratory (immortal-time bias)^a^ | ICU transfer after 24 hours, OR | 0.68 (0.46–1.00) |
|  |  | DT not present on admission, RR | 0.04 (0.01–0.18) |
|  |  | Length of stay, RGM | 0.78 (0.74–0.83) |
|  |  | 30-day death or return to hospital, OR | 0.94 (0.75–1.17) |
| **Exposure redefined as consultation within 48 hours** | | | |
|  |  | ICU transfer after 24 hours, OR | 0.63 (0.47–0.84) |
|  |  | DT not present on admission, RR | 0.47 (0.22–0.97) |
|  |  | Phenobarbital, any, OR | 1.16 (0.99–1.35) |
|  |  | Thiamine within 24 hours, OR | 1.22 (1.07–1.38) |
|  |  | Length of stay, RGM | 0.94 (0.90–0.99) |
|  |  | 30-day death or return to hospital, OR | 0.75 (0.65–0.88) |
| **1:1 propensity-matched cohort (n = 2,646)** | | | |
|  |  | ICU transfer after 24 hours, OR | 0.83 (0.59–1.17) |
|  |  | DT not present on admission, OR | 0.17 (0.04–0.74) |
|  |  | Phenobarbital, any, OR | 1.22 (1.02–1.46) |
|  |  | Thiamine within 24 hours, OR | 1.24 (1.06–1.45) |
|  |  | 30-day death or return to hospital, OR | 0.83 (0.69–0.98) |
| **Additional adjustment for palliative care consultation within 24 hours** | | | |
|  |  | ICU transfer after 24 hours, OR | 0.73 (0.54–0.99) |
|  |  | DT not present on admission, RR | 0.08 (0.02–0.32) |
|  |  | In-hospital death, OR | 0.41 (0.20–0.83) |
|  |  | 30-day death or return to hospital, OR | 0.78 (0.66–0.91) |
| **E-values (point estimate; CI limit)** | | | |
|  |  | ICU transfer after 24 hours | 2.1; 1.1 |
|  |  | 30-day death or return to hospital | 1.5; 1.3 |

^a^A later consult requires remaining in hospital beyond 24 hours and may be prompted by deterioration, so these comparisons are biased toward later consultation. ^b^Palliative care consultation within 24 hours; mechanical ventilation and code-status orders were not retrievable.. Abbreviations: DT, delirium tremens; eq, equivalent; OR, odds ratio; RD, overlap-weighted risk difference; RGM, ratio of geometric means; RR, overlap-weighted risk ratio.

**Supplementary Table 3. STROBE Statement Checklist**

|  | **Item No.** | **Recommendation** | **Page/Section** | **Relevant text from manuscript** |
| --- | --- | --- | --- | --- |
| Title and abstract | 1a  1b | Indicate the study’s design with a commonly used term in the title or the abstract | ABSTRACT; Methods | **Abstract:** Retrospective single-center cohort of adult hospitalizations with alcohol withdrawal (diagnosis code or CIWA-Ar ≥10 within 24 hours) from July 2020 to January 2026. |
|  |  | Provide in the abstract an informative and balanced summary of what was done and what was found | ABSTRACT | Of 4,663 hospitalizations, 1,379 (29.6%) received early consultation. After adjustment, early consultation was associated with lower odds of ICU transfer (OR 0.73; 95% CI 0.54–0.99) and fewer episodes of delirium tremens (RD −1.09 per 100; −1.45 to −0.73), with no significant difference in withdrawal seizure. Early consultation was associated with phenobarbital and thiamine within 24 hours, higher 48-hour lorazepam-equivalent dose, shorter stays, and lower odds of 30-day death or return to hospital. Conclusions: … lower odds of ICU transfer, and fewer episodes of delirium tremens, although the complication associations were sensitive to cohort definition. |
| **Introduction** | | | |  |
| **Background/rationale** | 2 | Explain the scientific background and rationale for the investigation being reported | INTRODUCTION | Alcohol withdrawal complicates more than 500,000 hospitalizations annually in the United States and can cause seizures, delirium tremens, and intensive care unit (ICU) admission. Guidelines recommend symptom-triggered benzodiazepine therapy, early thiamine, and structured monitoring, yet inpatient care is characterized by variable benzodiazepine use, delayed thiamine, and limited uptake of medications for alcohol use disorder… Prior evaluations rarely examined withdrawal-specific processes such as benzodiazepine dosing, phenobarbital use, and thiamine timing, and most defined exposure as consultation at any point during the stay. Whether ACS changes withdrawal-specific care and short-term outcomes, and whether the timing of consultation matters, remains uncertain. |
| **Objectives** | 3 | State specific objectives, including any prespecified hypotheses | INTRODUCTION | We hypothesized that management of acute withdrawal during the first day is a principal pathway, and examined whether ACS consultation within 24 hours of arrival was associated with withdrawal pharmacotherapy and in-hospital complications, comparing early with later consultation to separate timing from consultation itself. |
| **Methods** | | | |  |
| **Study design** | 4 | Present key elements of study design early in the paper | METHODS; Study Design and Setting | We conducted a retrospective cohort study at Vanderbilt University Hospital, using electronic health record data from January 2018 to January 2026 |
| **Setting** | 5 | Describe the setting, locations, and relevant dates, including periods of recruitment, exposure, follow-up, and data collection | METHODS; Study Design and Setting; Participants | Vanderbilt University Hospital; electronic health record data from January 2018 to January 2026. The ACS is a multidisciplinary team of addiction physicians, advanced practice clinicians, and social workers that provides daytime consultation seven days a week to all inpatient services; the service was formally staffed and promoted to admitting teams in January 2021. Throughout the study period, withdrawal management on all services followed an institutional order set aligned with national guidelines. The primary cohort comprised confirmed withdrawal hospitalizations in the steady-state period (July 2020 to January 2026); exposure was assessed within 24 hours of arrival and outcomes during the hospitalization and within 30 days of discharge. |
| Participants | 6a | Cohort study—Give the eligibility criteria, and the sources and methods of selection of participants. Describe methods of follow-up | METHODS; Participants | We identified adult (≥18 years) inpatient hospitalizations with evidence of alcohol withdrawal, defined as either an alcohol withdrawal diagnosis (ICD-10, F10.13x or F10.23x) or a maximum CIWA-Ar score of 10 or higher within 24 hours of arrival. We excluded hospitalizations shorter than 24 hours, deaths within 24 hours, and admissions to the psychiatric hospital… The primary cohort comprised confirmed withdrawal hospitalizations in the steady-state period (July 2020 to January 2026). Hospitalizations with lower CIWA-Ar scores or before July 2020 were included in sensitivity analyses. Thirty-day outcomes were assessed among patients discharged alive. |
|  | 6b | Cohort study—For matched studies, give matching criteria and number of exposed and unexposed | METHODS; other analyses | The primary analysis used overlap weighting rather than matching. As a sensitivity analysis we repeated the core analyses in a 1:1 propensity-matched cohort (caliper 0.2 SD of the logit); matched numbers are given in Supplementary Table 2. |
| **Variables** | 7 | Clearly define all outcomes, exposures, predictors, potential confounders, and effect modifiers. Give diagnostic criteria, if applicable | METHODS; Exposure, Outcomes, Covariates | **Exposure**  Exposure was an ACS consult order placed within 24 hours of time zero (early consultation)… The comparison group comprised hospitalizations with no or later consultation; the latter were examined separately in a timing analysis.  **Outcomes**  Our primary outcomes were withdrawal-specific in-hospital complications: (1) ICU transfer from a non-ICU service after a 24-hour landmark, (2) delirium tremens (F10.231 or F10.131) not present on admission, and (3) withdrawal seizure (R56.9 or G40.5x) not present on admission… Process outcomes were withdrawal pharmacotherapy… any benzodiazepine; lorazepam-equivalent dose…; phenobarbital and thiamine, at any time and within 24 hours; and adjunctive agents. Secondary outcomes were length of stay; discharge disposition; all-cause readmission, ED visit, and death within 30 days of discharge, and their composite, among patients discharged alive; and inpatient MAUD initiation and new MAUD at discharge.  **Covariates and effect modifiers**  Covariates, measured at or before hospitalization except CIWA-Ar variables, were age, sex, race, Hispanic ethnicity, and insurance; initial CIWA-Ar score, maximum score within 24 hours, and absence of any score during the stay; delirium tremens and seizure present on admission; admission from the ED to an ICU; prior delirium tremens, withdrawal seizure, and withdrawal hospitalization; comorbid conditions and the Elixhauser comorbidity count; prior MAUD, admissions and ED visits; admission year and weekend admission. Subgroups: withdrawal severity, prior utilization, and opioid use disorder. |
| **Data sources/ measurement** | 8* | For each variable of interest, give sources of data and details of methods of assessment (measurement). Describe comparability of assessment methods if there is more than one group | METHODS; Exposure, Outcomes, Covariates | All variables were derived from the electronic health record and assessed identically in exposed and unexposed hospitalizations. Exposure: consult order time (we used the order time because documentation of consult completion lags the clinical encounter); time zero was ED arrival for patients admitted through the ED so that ED medications and CIWA-Ar scores were attributed to the hospitalization. Complications: diagnosis codes with present-on-admission indicators (diagnoses with a present-on-admission indicator of yes, or recorded only on the ED or encounter diagnosis list, were classified as present on admission; undetermined or exempt indicators counted as neither). Process outcomes: medication administration records after time zero, with benzodiazepines converted to lorazepam equivalents. Covariates: diagnosis codes, flowsheet CIWA-Ar scores, medication lists, and utilization in the prior 12 months. |
| **Bias** | 9 | Describe any efforts to address potential sources of bias | METHODS | Time zero defined as ED arrival so that pre-admission care was attributed to the hospitalization; exposure defined by order time. ICU transfer analyzed from a 24-hour landmark to exclude transfers within the exposure window that preclude consultation rather than result from it. Delirium tremens and seizure present on admission treated as baseline covariates because they precede and commonly prompt consultation. Confounding addressed with propensity score overlap weighting on all covariates, additional outcome-model adjustment, and standard errors clustered on patient. Because 24-hour process outcomes could precede the consult order, we described time to the order and the proportion of first phenobarbital and thiamine doses given before it. E-values, an early-acuity sensitivity analysis, restriction to CIWA-Ar-confirmed entry, and omission of the within-window CIWA-Ar maximum addressed residual confounding and selection. |
| **Study size** | 10 | Explain how the study size was arrived at | METHODS; RESULTS | No formal sample size calculation was performed; the study included all eligible hospitalizations in the steady-state period (July 2020 to January 2026): 4,663 hospitalizations of 3,365 patients. |
| **Quantitative variables** | 11 | Explain how quantitative variables were handled in the analyses. If applicable, describe which groupings were chosen and why | METHODS; Statistical Analysis | CIWA-Ar scores (initial and maximum within 24 hours) were analyzed as continuous covariates, with missing scores set to zero with an indicator. Benzodiazepine dose was analyzed as a two-part model: any administration, then log-transformed lorazepam-equivalent dose among recipients, reported as ratios of geometric means. Length of stay was log-transformed. Subgroup categories: maximum CIWA-Ar within 24 hours <10, 10–18, ≥19; prior utilization ≥3 ED visits or ≥2 admissions in 12 months. |
| Statistical methods | 12a | Describe all statistical methods, including those used to control for confounding | METHODS; Statistical Analysis | We estimated a propensity score for early consultation by logistic regression on all covariates and applied overlap weighting… Because patients could contribute more than one hospitalization, standard errors were clustered on patient. We re-estimated weights in each analytic subset… We fit overlap-weighted logistic regression adjusting for withdrawal severity, admission from the ED to ICU, complications present on admission, prior withdrawal history, Elixhauser count, liver disease, opioid use disorder, and prior utilization, and report odds ratios (ORs) with 95% CIs; we also fit a cause-specific Cox model from the landmark with discharge as a competing event. For delirium tremens and seizure not present on admission, which had fewer than 10 exposed events, we report the overlap-weighted risk difference (RD) with patient-clustered delta-method CIs, without an outcome model. Binary process outcomes used the same overlap-weighted logistic models… log-transformed length of stay with overlap-weighted linear models; discharge disposition with cause-specific Cox models… 30-day and MAUD outcomes with overlap-weighted logistic models among patients discharged alive. |
|  | 12b | Describe any methods used to examine subgroups and interactions | METHODS | Third, we re-estimated weights within subgroups defined by withdrawal severity (maximum CIWA-Ar within 24 hours <10, 10–18, ≥19), prior utilization (≥3 ED visits or ≥2 admissions in 12 months), and opioid use disorder, without tests of interaction, and described outcomes stratified by PDD. |
|  | 12c | Explain how missing data were addressed | METHODS; DISCUSSION | Missing CIWA-Ar scores were set to zero with a missingness indicator. Covariates were complete except insurance; hospitalizations without a coverage record (8%) were classified as self-pay/other. |
|  | 12d | Cohort study—If applicable, explain how loss to follow-up was addressed |  | In-hospital outcomes were complete by design. Thirty-day outcomes were assessed among patients discharged alive using health-system records; readmissions and deaths outside the health system were not captured (Limitations). |
|  | 12e | Describe any sensitivity analyses | METHODS; RESULTS | We performed three sets of additional analyses. First, … we compared early with later consultation among consulted hospitalizations… Second, we repeated the core analyses in six alternative cohorts (Supplementary Table 1); with exposure redefined as consultation within 48 hours; in a 1:1 propensity-matched cohort; with adjustment for palliative care consultation within 24 hours as an acuity marker; among hospitalizations without delirium tremens or seizure at admission; with entry restricted to a maximum CIWA-Ar of 10 or higher within 24 hours; and omitting the 24-hour maximum CIWA-Ar. We calculated E-values for the primary outcomes. Third, … subgroups… |
| **Results** | | | |  |
| Participants | 13a* | Report numbers of individuals at each stage of study—eg numbers potentially eligible, examined for eligibility, confirmed eligible, included in the study, completing follow-up, and analysed | RESULTS | Among 12,913 hospitalizations with evidence of alcohol withdrawal over the full study period, 8,796 (68%) were identified by CIWA-Ar monitoring alone… The primary cohort comprised 4,663 hospitalizations of 3,365 patients, of which 1,379 (29.6%) received ACS consultation within 24 hours and 3,284 did not, including 832 consulted later in the stay (Figure 2). Among patients discharged alive (n = 4,538), 30-day outcomes were assessed. |
|  | 13b | Give reasons for non-participation at each stage | METHODS | We excluded hospitalizations shorter than 24 hours, deaths within 24 hours, and admissions to the psychiatric hospital. Hospitalizations with lower CIWA-Ar scores or before July 2020 were included in sensitivity analyses (Figure 2). |
|  | 13c | Consider use of a flow diagram | FIGURE 2 | Figure 2. Study flow diagram. |
| Descriptive data | 14a* | Give characteristics of study participants (eg demographic, clinical, social) and information on exposures and potential confounders | RESULTS | Patients with early ACS were younger and more often had severe withdrawal, complications present on admission, prior withdrawal hospitalization, and opioid use disorder; after overlap weighting, all covariates were balanced (effective sample size 2,354 unexposed, 1,267 exposed; Table 1). |
|  | 14b | Indicate number of participants with missing data for each variable of interest | METHODS; DISCUSSION | Covariates were complete except insurance; hospitalizations without a coverage record (8%) were classified as self-pay/other. CIWA-Ar monitoring is not applied uniformly across services (16% of unexposed hospitalizations had no score within 24 hours); missing scores were set to zero with a missingness indicator. |
|  | 14c | Cohort study—Summarize follow-up time (eg, average and total amount) | METHODS | Follow-up comprised the index hospitalization (length of stay, median [IQR], by group in Table 2) and 30 days after discharge for patients discharged alive. |
| **Outcome data** | 15* | Cohort study—Report numbers of outcome events or summary measures over time | RESULTS | Early ACS hospitalizations less often had ICU transfer after 24 hours (6.5% vs 8.5%), delirium tremens not present on admission (2 [0.1%] vs 64 [1.9%]), and withdrawal seizure not present on admission (4 [0.3%] vs 27 [0.8%]), but more often had these complications present on admission; 352 of 430 coded delirium tremens hospitalizations (82%) were present on admission (Table 2). They received higher 48-hour lorazepam-equivalent doses (median 10 vs 4 mg), more phenobarbital and thiamine within 24 hours, and less dexmedetomidine and midazolam, and had shorter stays, lower mortality, and more PDD and MAUD initiation. |
| Main results | 16a | Give unadjusted estimates and, if applicable, confounder-adjusted estimates and their precision (eg, 95% confidence interval). Make clear which confounders were adjusted for and why they were included | RESULTS, Unadjusted and Adjusted Outcomes; Tables 2–3; METHODS, Statistical Analysis | Unadjusted: Table 2. Adjusted: In weighted models, early ACS consultation was associated with lower odds of ICU transfer after 24 hours (OR 0.73; 95% CI 0.54–0.99; cause-specific HR 0.76; 0.57–1.01), fewer episodes of delirium tremens not present on admission (RD −1.09 per 100 hospitalizations; −1.45 to −0.73), and a nonsignificant reduction in withdrawal seizure not present on admission (RD −0.34 per 100; −0.73 to 0.05) (Table 3)… higher lorazepam-equivalent dose in the first 48 hours (RGM 1.44; 1.32–1.57)… phenobarbital within 24 hours (OR 1.57; 1.32–1.86) and thiamine within 24 hours (OR 1.23; 1.07–1.41)… shorter length of stay (RGM 0.92; 0.88–0.97)… lower odds of the 30-day composite (OR 0.78; 0.66–0.91). Confounders adjusted for are listed in Methods (propensity model on all covariates; outcome models adjusted for withdrawal severity, admission from the ED to ICU, complications present on admission, prior withdrawal history, Elixhauser count, liver disease, opioid use disorder, and prior utilization). |
|  | 16b | Report category boundaries when continuous variables were categorized | METHODS | Subgroups defined by withdrawal severity (maximum CIWA-Ar within 24 hours <10, 10–18, ≥19) and prior utilization (≥3 ED visits or ≥2 admissions in 12 months). |
|  | 16c | If relevant, consider translating estimates of relative risk into absolute risk for a meaningful time period | RESULTS | Rare complications are reported as overlap-weighted absolute risk differences per 100 hospitalizations (delirium tremens RD −1.09 per 100; withdrawal seizure RD −0.34 per 100), with unadjusted proportions in Table 2. |
| **Other analyses** | 17 | Report other analyses done—eg analyses of subgroups and interactions, and sensitivity analyses | RESULTS; Additional Analyses | Among 2,211 hospitalizations consulted at any time, early compared with later consultation was associated with phenobarbital within 24 hours (OR 1.61; 1.26–2.06), higher 48-hour lorazepam-equivalent dose (RGM 1.29; 1.15–1.46), and thiamine within 24 hours (OR 1.25; 1.04–1.51)… Among early consultations, the order was placed a median of 14.1 hours after time zero, and 60.5% of first phenobarbital and 69.3% of first thiamine doses within 24 hours preceded it… Estimates were similar with a 48-hour exposure window, in the matched cohort, with adjustment for palliative care consultation, among hospitalizations without delirium tremens at admission, and without the 24-hour maximum CIWA-Ar; E-values were 2.1 for ICU transfer and 1.5 for the 30-day composite (Supplementary Table 2). Restricting entry to a maximum CIWA-Ar of 10 or higher within 24 hours… attenuated ICU transfer (OR 0.82; 0.58–1.15) and delirium tremens (RR 0.24; 0.05–1.08). Phenobarbital associations were larger in severe withdrawal and among higher utilizers… PDD hospitalizations… |
| **Discussion** | | | |  |
| **Key results** | 18 | Summarise key results with reference to study objectives | DISCUSSION | Among adults hospitalized with alcohol withdrawal, ACS consultation within 24 hours of arrival was associated with earlier and more intensive withdrawal treatment: more phenobarbital, higher 48-hour benzodiazepine dosing, earlier thiamine, and less dexmedetomidine. Early consultation was also associated with lower odds of ICU transfer after the first day and fewer episodes of delirium tremens arising after admission. Process, length-of-stay, and 30-day findings were robust across sensitivity analyses, whereas the ICU transfer and delirium tremens associations were attenuated and no longer significant when cohort entry required CIWA-Ar confirmation. |
| **Limitations** | 19 | Discuss limitations of the study, taking into account sources of potential bias or imprecision. Discuss both direction and magnitude of any potential bias | DISCUSSION, Limitations | This single-center study reflects one service, its referral patterns, and local withdrawal protocols. Cohort entry relied on CIWA-Ar monitoring… and on diagnosis codes assigned at discharge; hospitalizations entered by code alone were predominantly unexposed and may include patients too ill to be scored or consulted, and without them the ICU transfer and delirium tremens associations were no longer statistically significant. As noted, the 24-hour process associations partly reflect admitting-team treatment that preceded the consult order. Residual confounding by clinician selection remains possible: E-values indicate that an unmeasured confounder associated with exposure and outcome by a risk ratio of about 2 could explain the ICU transfer association… Delirium tremens and seizure were ascertained from diagnosis codes, and present-on-admission assignment depends on documentation… inflating the association; this cannot be excluded without chart review. Subgroup differences were not formally tested. Readmissions and deaths outside the health system were not captured. Process measures describe the intensity and timing of therapy, not its appropriateness, and we did not assess oversedation or other adverse effects. |
| **Interpretation** | 20 | Give a cautious overall interpretation of results considering objectives, limitations, multiplicity of analyses, results from similar studies, and other relevant evidence | DISCUSSION, CONCLUSION | Withdrawal management is a plausible pathway, although the evidence here is indirect… Two findings nonetheless point to consultation itself: higher benzodiazepine dosing over 48 hours, which extends past most consult orders, and the early-versus-later comparison… Conclusion: Among hospitalized adults with alcohol withdrawal, addiction consultation within 24 hours of arrival was associated with earlier, more intensive withdrawal pharmacotherapy, shorter hospitalizations, lower odds of ICU transfer, and fewer episodes of delirium tremens, although the complication associations were sensitive to cohort definition. Withdrawal management during the first day is a plausible, though not fully isolated, pathway for the short-term benefits of consultation that discharge pharmacotherapy does not explain. |
| **Generalisability** | 21 | Discuss the generalisability (external validity) of the study results | DISCUSSION, Limitations | This single-center study reflects one service, its referral patterns, and local withdrawal protocols. |
| **Other information** | | | |  |
| **Funding** | 22 | Give the source of funding and the role of the funders for the present study and, if applicable, for the original study on which the present article is based | ACKNOWLEDGEMENTS, Funders | Agency for Healthcare Research and Quality (AHRQ) (K08HS029695; principal investigator, T.J.R.). The funder had no role in the design, conduct, analysis, or reporting of the study. |
